# Nucleocapsid antigenemia is a marker of acute SARS-CoV-2 infection

**DOI:** 10.1101/2022.01.23.22269354

**Authors:** Hans P. Verkerke, Gregory L. Damhorst, Daniel S. Graciaa, Kaleb McLendon, William O’Sick, Chad Robichaux, Narayanaiah Cheedarla, Sindhu Potlapalli, Shang-Chuen Wu, Kristin R.V. Harrington, Andrew Webster, Colleen Kraft, Christina A. Rostad, Jesse J. Waggoner, Neel R. Gandhi, Jeannette Guarner, Sara C. Auld, Andrew Neish, John D. Roback, Wilbur A. Lam, N. Sarita Shah, Sean R. Stowell

**Author notes:** Corresponding authors: Sean Stowell, MD, PhD, Brigham and Women’s Hospital, Harvard Medical School, Sarita Shah, MD, MPH, Emory University Rollins School of Public Health. Equal contribution (Hans P. Verkerke, Gregory L. Damhorst, N. Sarita Shah, Sean R. Stowell).

## Abstract

**Background:** Reliable detection of SARS-CoV-2 infection is essential for diagnosis and treatment of disease as well as infection control and prevention during the ongoing COVID-19 pandemic. Existing nucleic acid tests do not reliably distinguish acute from resolved infection, as residual RNA is frequently detected in the absence of replication-competent virus. We hypothesized that viral nucleocapsid in serum or plasma may be a specific biomarker of acute infection that could enhance isolation and treatment strategies at an individualized level.

**Methods:** Samples were obtained from a retrospective serological survey using a convenience sampling method from adult inpatient and outpatient encounters from January through March 2021. Samples were categorized along a timeline of infection (e.g. acute, late presenting, convalescent) based on timing of available SARS-CoV-2 testing and symptomatology. Nucleocapsid was quantified by digital immunoassay on the Quanterix HD-X platform.

**Results:** In a large sample of 1860 specimens from 1607 patients, the highest level and frequency of antigenemia were observed in samples obtained during acute SARS-CoV-2 infection. Levels of antigenemia were highest in samples from seronegative individuals and in those with more severe disease. Using ROC analysis, we found that antigenemia exhibited up to 85.8% sensitivity and 98.6% specificity as a biomarker for acute COVID-19.

**Conclusions:** Nucleocapsid antigenemia is a sensitive and specific biomarker for acute SARS-CoV-2 infection and may aid in individualized assessment of SARS-CoV-2 infection resolution or persistence, although interpretation is limited by absence of a diagnostic gold standard for active infection.

## Introduction

Although the standard of care for SARS-CoV-2, reverse transcription polymerase chain reaction (RT-PCR) remains an imperfect diagnostic marker for coronavirus disease 2019 (COVID-19) because SARS-CoV-2 RNA commonly persists beyond the period of acute infection [1-3]. Accordingly, Centers for Disease Control and Prevention (CDC) guidelines do not recommend re-testing most individuals by RT-PCR within 90 days following diagnosis. Instead, isolation guidelines are based on time from symptom onset [4, 5]. This creates a dilemma when screening tests detect SARS-CoV-2 RNA in a patient without well-defined onset or resolution of COVID-like illness. Alternative molecular markers for acute infection are not widely available [6] and low sensitivity respiratory antigen testing may be effectively applied at a population level [7, 8], but there remains a need for more sensitive and specific diagnostics to provide individualized guidance.

The presence of viral nucleocapsid protein in peripheral blood (antigenemia) has been demonstrated in SARS-CoV-1 and SARS-CoV-2 infection [9-21]. A blood-based antigen biomarker may have inherent advantages over upper respiratory tract antigen testing, or biomarkers such as RT-PCR cycle threshold (Ct) value and sub-genomic RNA (sgRNA), because specimen quality and quantity can be standardized. Reports of antigenemia test performance as a diagnostic biomarker are inconsistent, likely due to varying assay composition and inconsistent reference standards as many studies compare against respiratory RT-PCR as a gold standard and fail to account for the persistence of RNA beyond acute infection.

In this study, we present evidence from a large serosurvey of adults in inpatient and outpatient settings to explore the hypothesis that nucleocapsid antigenemia is a sensitive and specific marker of acute infection as defined by a clinical timeline. Specifically, each blood sample was categorized through rigorous review of clinical history and respiratory SARS-CoV-2 testing in a schema that assumes a typical course of COVID-19 for all subjects. We find a strong association between acute infection and nucleocapsid antigenemia, which also correlates with serostatus and disease severity. Together our findings suggest antigenemia may clarify disease timing and provide needed insight in many clinical settings.

## Methods

### Clinical specimens

We collected a convenience sample of residual plasma, serum, and whole blood specimens from the clinical chemistry laboratory of Emory Medical Laboratories one day per week between January 11, 2021 and March 12, 2021. These specimens were originally collected for routine clinical testing from inpatient (medical/surgical wards, intensive care, obstetrics) and outpatient settings (clinics, emergency department, infusion centers, ambulatory surgery). Samples were transferred to a –80C repository after clinical testing was completed, but prior to being discarded. More than one blood sample from the same patient was permitted with a minimum time of five days between samples. This study was approved and granted complete HIPAA and consent waiver by the Emory University Institutional Review Board (STUDY00000510).

### Nucleocapsid assay

Nucleocapsid antigenemia was quantified on the Quanterix HD-X platform. Residual serum and plasma samples were thawed once after storage at –80°C and diluted 1 to 3 in assay sample diluent. Diluted samples were then run using the ultrasensitive SIMOA SARS-CoV-2 N Protein Antigen assay on the automated Quanterix HD-X platform (Quanterix, Billerica, MA, USA) which has a validated limit of detection of 0.099 pg/mL in respiratory and saliva samples. Samples with antigen levels too high for the linear range of the assay were further diluted 1 to 20 and re-tested. Final antigen concentrations were determined by interpolation after sigmoidal fitting of duplicate calibration curves run on each test plate.

### Serological Testing

In-house developed single-dilution serological screening assays for SARS-CoV-2 receptor binding domain (RBD) and nucleocapsid antibodies were used to establish serological status at the time of antigenemia testing. Antibody class-specific RBD serologies were performed as previously described [22]. Nucleocapsid antibody testing was performed using an in-house developed ELISA (supplementary information).

### Medical record review

Patient medical record number was recorded at the time of specimen collection. The Emory Healthcare Clinical Data Warehouse (CDW) was queried for SARS-CoV-2 nucleic acid amplification tests (NAAT), clinical notes, ICD-10 codes, laboratory values, mechanical ventilation, and date of death. All Ct values were obtained directly from reports produced by the manufacturer’s software (supplementary information).

A COVID-19 status label (positive or negative) and a category (convalescent, late-presenting, acute, pre-COVID, and same-day negative) were assigned to each blood sample based on that patient’s (1) SARS-CoV-2 respiratory testing (including NAAT or antigen), (2) date of earliest positive test, and (3) date of symptom onset (**Figure 1**).

**Figure 1.**
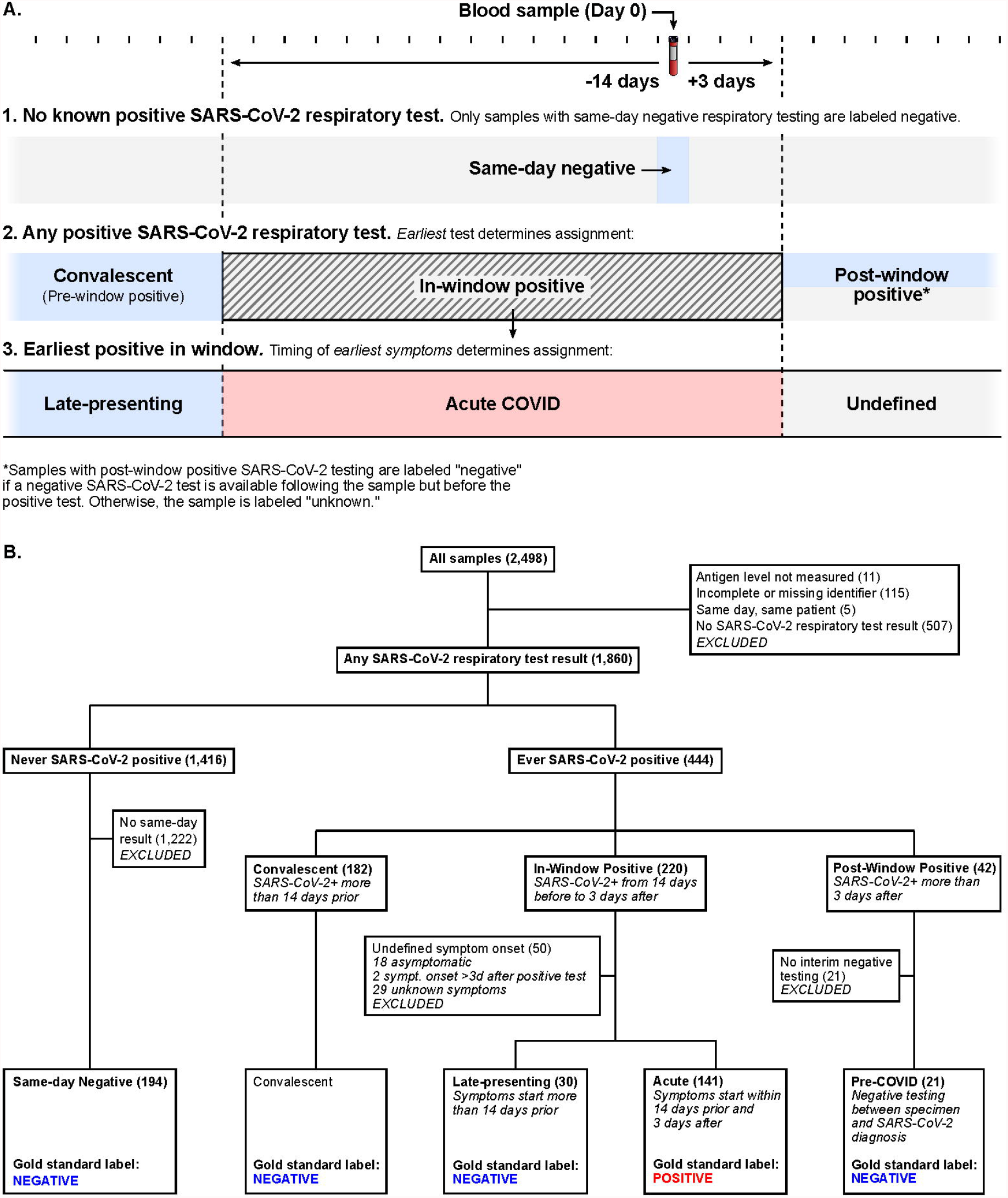
(A) Schematic of process for COVID status assignment. Samples from patients with no record of positive SARS-CoV-2 respiratory testing were only considered negative if corresponding negative respiratory testing occurred on the same day. Due to the lack of a gold standard for active SARS-CoV-2 infection, samples from individuals with history of positive SARS-CoV-2 testing are labeled based on earliest known positive SARS-CoV-2 respiratory test and time since symptom onset. (B) Flow chart of categorization and labeling process indicating number of samples assigned to each group.

Chart review began with automated review of NAAT results available in the medical record. Blood samples from a patient with a positive NAAT more than fourteen days prior to sample collection were labeled *convalescent* and no further review of the medical record for categorization purposes was performed. History and physical clinical notes dated within fourteen days before or after the date of the blood sample were then reviewed, if available, for all patients not labeled *convalescent*. Date of COVID-like symptom onset (including fever, fatigue, malaise, myalgia, headache, dyspnea, cough, wheezing, anosmia, ageusia, congestion, rhinorrhea, or diarrhea) and earliest positive SARS-CoV-2 testing (NAAT or antigen) was recorded if these had been described in the history narrative or clinician’s assessment and plan.

The original medical records were then reviewed for all patients (other than those labeled *convalescent*) with a positive SARS-CoV-2 test who did not yet have date of symptom onset recorded in our data set. The entire medical record was available during this stage, but the reviewer was blinded to antigenemia status which was not considered in labeling of COVID-19 status or category assignment.

Given that re-infection with SARS-CoV-2 was rare at the time of this study [23], our approach assumed that no re-infection events were captured in our sample set, which spanned 3 months. Patients without any record of SARS-CoV-2 testing were excluded from analysis. Further detail is provided in Supplementary Information.

### Data analysis

Data obtained during specimen collection were stored in Microsoft Excel. CDW reports were provided in .csv format. All data were then imported into MATLAB (The MathWorks, Inc.) for analysis. Wilcoxon rank-sum test was used for comparisons.

## Results

### Specimens & COVID-19 status assignments

2,498 serum and plasma samples were targeted for evaluation during the study period (**Figure 1B**). Eleven samples were not evaluated for antigenemia due to pre-analytical factors such as insufficient sample volume. 2,487 samples were available for quantification of antigenemia, of which 255 (10.2%) exhibited detectable nucleocapsid. 115 of 2,487 were excluded due to lack of patient identifiers and five additional samples were excluded as they had been collected on the same day as another blood sample from a single patient. Clinical data were examined for the remaining 2,367 samples from 2,101 unique patients (**Table 1**). 507 of 2,367 samples were excluded because of no record of SARS-CoV-2 testing, and 11 of these 507 (2.1%) had detectable antigenemia.

**Table 1.** Summary of patient characteristics by COVID status. *Reflects all included samples (including multiple samples for a unique patient). CDU = clinical decision unit

|  | Positive | Negative | Undefined |
| --- | --- | --- | --- |
| **N** | 130 | 385 | 1622 |
| **Age** mean (IQR) | 60.6 (52.2–73.0) | 54.2 (39.2–69.6) | 55.0 (39.8–70.2) |
| **Female** % | 47.7 | 57.1 | 55.5 |
| **Vaccinated** % | 1.5 | 4.7 | 9.9 |
| **Race** % |  |  |  |
| African American or Black | 78.5 | 72.0 | 60.7 |
| American Indian or Alaskan Native | 0.0 | 0.0 | 0.2 |
| Asian | 0.8 | 1.0 | 1.6 |
| Caucasian or White | 13.1 | 20.0 | 29.0 |
| Native Hawaiian or Other Pacific Islander | 0.0 | 0.3 | 0.2 |
| Multiple | 0.0 | 0.5 | 0.4 |
| Unknown, Unavailable or Unreported | 7.7 | 6.2 | 7.9 |
| **Ethnicity** |  |  |  |
| Non-Hispanic or Latino | 83.9 | 86.0 | 84.4 |
| Unreported, Unknown, Unavailable | 13.9 | 7.8 | 11.9 |
| Hispanic or Latino | 0.8 | 5.5 | 3.0 |
| Not Recorded | 1.5 | 0.8 | 0.7 |
| **Antigenemia\*** % | 85.8 | 10.1 | 3.9 |
| **Setting\*** |  |  |  |
| Inpatient | 70.9 | 42.9 | 39.3 |
| ER or CDU | 29.1 | 27.2 | 14.1 |
| Outpatient | 0.0 | 26.7 | 45.3 |
| Peripartum | 0.0 | 3.3 | 1.1 |

The remaining 1,860 samples from 1,607 patients had SARS-CoV-2 testing records to guide categorization and were classified as described in **Figure 1B** and **Table 2**.

**Table 2.** Categories determined by chart review for samples and patients included in the analysis. NAAT = Nucleic acid amplification testing; *Includes in-hospital NAAT as well as community NAAT or antigen testing if reported in the clinical narrative

|  | Samples | Unique patients |
| --- | --- | --- |
| **Never SARS-CoV-2 positive** | 1416 | 1249 |
| Same-day negative NAAT | 194 | 194 |
| **Ever SARS-CoV-2 positive\*** | 444 | 360 |
| Convalescent | 182 | 153 |
| Late-presenting | 30 | 30 |
| Acute | 141 | 130 |
| Sampled three or more days prior to diagnosis | 42 | 34 |
| Negative interim testing | 21 | 16 |

### Diagnostic performance of antigenemia for acute COVID-19

Nucleocapsid antigenemia was present at higher frequency and with a higher median concentration in acute COVID samples compared to samples categorized as late-presenting, convalescent, pre-COVID, or same-day negative (p < 0.001 for all comparisons; **Figure 2A**). ROC analysis demonstrated area under the curve (AUC) of 0.902 in distinguishing samples from patients experiencing acute infection from all non-acute categories, and sensitivity and specificity were 85.2% and 89.9%, respectively (**Figure 2B**). Test characteristics with censoring of the potentially ambiguous late-presenting group showed AUC 0.914, sensitivity 85.8%, and specificity 93.7% while the most stringent comparison (censoring of the convalescent and late-presenting groups) demonstrated AUC 0.972, sensitivity 85.8%, and specificity 98.6%. Sensitivity improved to 93.9% when the comparison was only made among seronegative individuals (**Supplementary Figure 1**).

**Figure 2.**
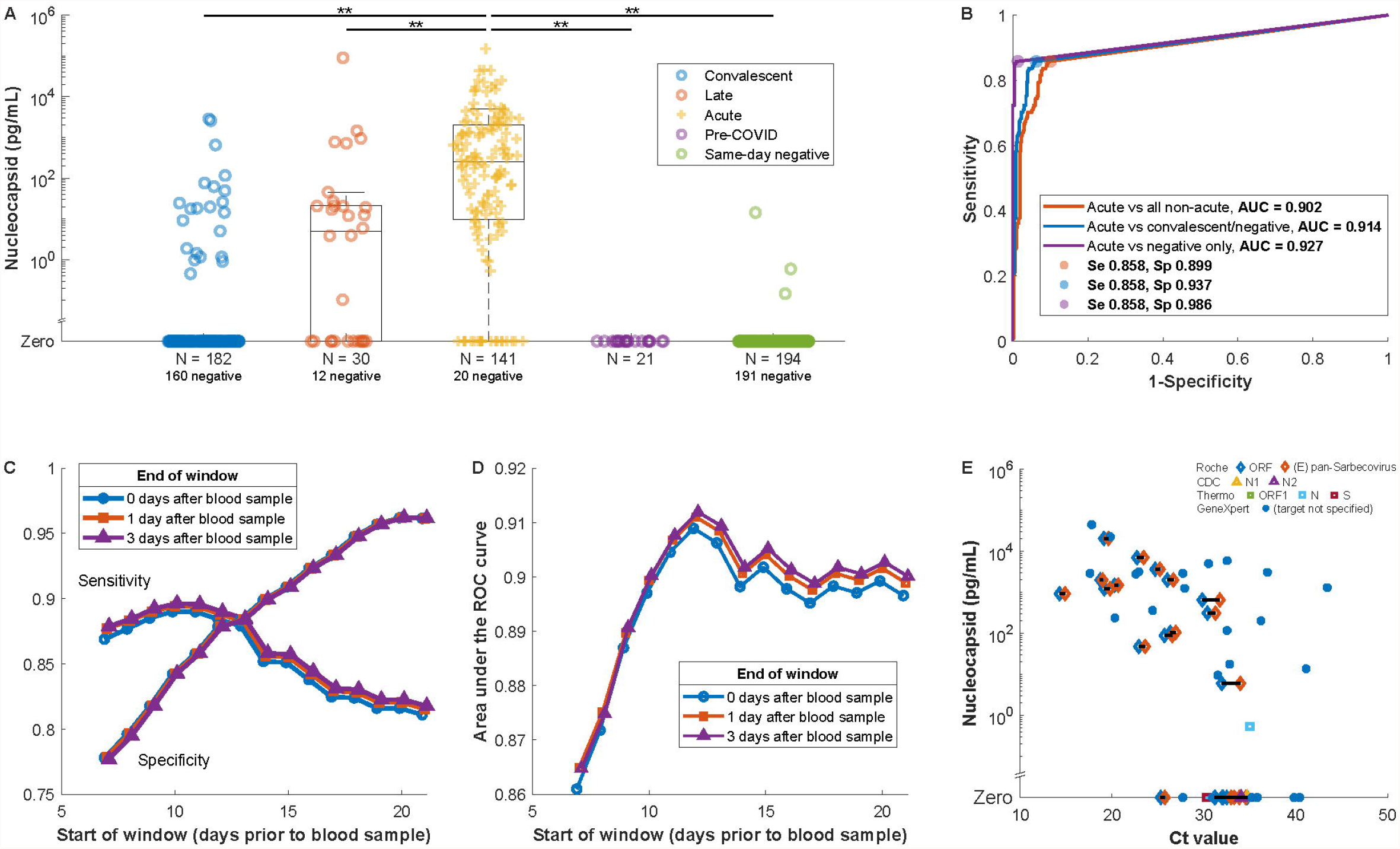
(A) Prevalence of antigenemia and serum or plasma nucleocapsid levels for blood samples by category. Unexpected results (presence of nucleocapsid in the convalescent and same-day negative groups, absence of nucleocapsid in the acute group) are examined in Supplementary Information Tables 2-5. (B) ROC curve for diagnostic performance of detectable antigenemia with reference to a -14/+3 day window for acute infection. The additional curves progressively exclude ambiguous categories. (C) Impact on sensitivity and specificity of varying the window period, which defines the reference standard for acute COVID. (D) AUC for the same varied window periods. (E) Antigenemia compared to RTPCR Ct value for those specimens with a Ct value available from the clinical laboratory on the same day. Symbols correspond to assay and gene target with horizontal line linking Ct values for different targets detected in the same sample. This includes data from four assays on three thermocycler platforms described in further detail in Supplementary Information.

Test characteristics were also examined when adjusting the reference standard by varying parameters of the acuity window. Sensitivity decreased as the window start period increased beyond -11 days (**Figure 2C**). Meanwhile, specificity consistently increased as the period of the acuity window was lengthened. Maximum AUC was observed with a window period opening at -12 days (AUC = 0.912 with window close at +3 days) with minimal effect of varying the post-sampling period from 0 to +3 days (**Figure 2D**).

Ct values from positive nasopharyngeal RT-PCR were available from the same day as a blood sample for 49 specimens. Only 6 of 17 samples with corresponding to Ct values greater than 33 had antigenemia and four of six of these were from the GeneXpert assay (**Figure 2E**). All except for two samples with corresponding Ct values less than 30 exhibited antigenemia.

### Temporal trends in antigenemia

We analyzed the dynamics of antigen level over time in samples from the acute, late-presenting and convalescent groups. The frequency of detectable nucleocapsid and antigen concentration decreased over time following diagnosis and reported symptom onset **(Figures 3A-B**). 18 samples were identified from patients who were asymptomatic at the time of COVID-19 diagnosis, 5 (27.7%) of which had detectable nucleocapsid antigenemia. Nucleocapsid antigen was detected more frequently (50.0%) in the subset of samples available from asymptomatic patients within 3 days of their diagnosis **(Figure 3C**). Among 55 samples from individuals with positive respiratory RT-PCR testing on the same day, seven convalescent samples did not exhibit antigenemia (**Figure 3D, Supplementary Table 1**) and acute infections primarily exhibited high antigenemia. Among this subset of patients, no antigenemia was observed more than fourteen days after the earliest known positive test **(Figure 3D**).

**Figure 3.**
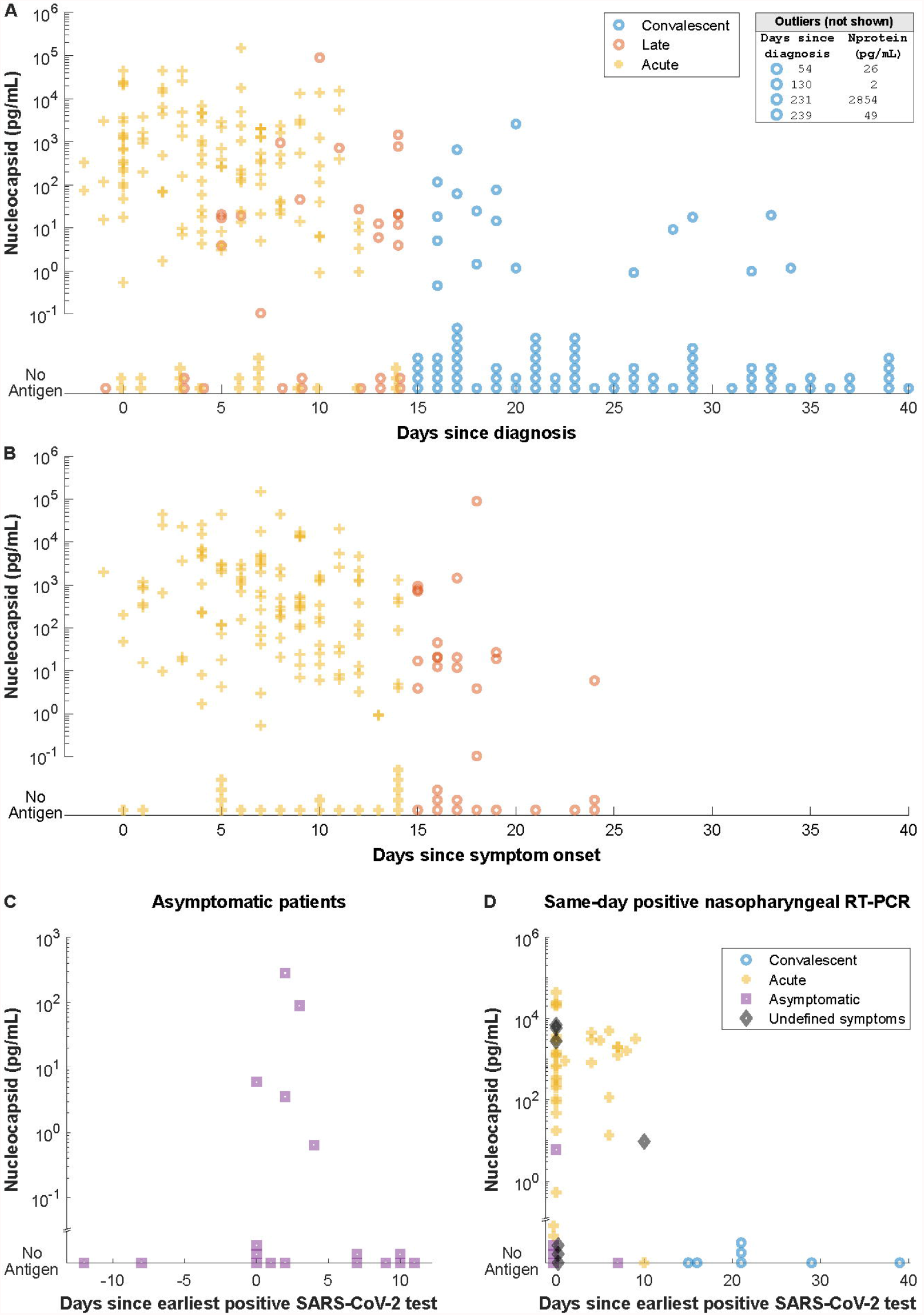
(A) Serum or plasma nucleocapsid plotted against time since diagnosis (top) and symptom onset (bottom, shown with an inverted y axis). Samples without antigen detected are shown stacked on the common horizontal axis. Four samples with antigenemia beyond 41 days are listed in the box and 93 samples without antigenemia between 41 and 351 days after earliest diagnosis are not shown. (B) Serum or plasma nucleocapsid in patients whose COVID-19 course was described as asymptomatic in clinical records. The x axis reflects time in between first known positive respiratory test and the day the blood sample used in our analysis was collected. (C) Serum or plasma nucleocapsid for individuals with positive nasopharyngeal RT-PCR on the same day as blood sample collection.

### Examination of outliers

We reviewed medical records for individuals with unexpected presence or absence of antigenemia. Twenty-one samples in the convalescent group had antigenemia (**Supplementary Table 2, Supplementary Figure 2**). Among these, two individuals had clinical history consistent with re-infection by SARS-CoV-2, two were highly immunocompromised, and eleven samples (median time from diagnosis 20 days, IQR 16.5-28.5 days) had severe COVID-19 marked by need for high-flow oxygen, intubation or death. End-stage renal disease or dialysis was more common among samples in the convalescent group with antigenemia compared to those without antigenemia (fraction [95% confidence interval] = 0.41 [0.20-0.61] vs 0.13 [0.07-0.18]) whereas other co-morbidities were not significantly different (**Supplementary Figure 3**). Three individuals had negative respiratory SARS-CoV-2 testing and antigenemia on the same day, none of which had evidence of COVID-related symptoms (**Supplementary Table 3**). Eighteen samples had antigenemia after more than fourteen days of symptoms, of which fourteen were seropositive for both N IgG and RBG IgG, two seropositive for RBD IgG only, and two were seronegative for both. Thirteen had nucleocapsid level less than 46 pg/mL while the other five exceeded 700 pg/mL including both N and RBD seronegative patients and N-/RBD+ sample (**Supplementary Table 4, Supplementary Figures 4 & 5**). Twenty individuals with samples categorized in the acute COVID group did not have antigenemia – ten of these were collected ten or more days after symptom onset (**Supplementary Table 5, Supplementary Figure 6**).

### Antigenemia trends by antibody serostatus

Distribution of nucleocapsid levels in the acute COVID group were significantly different with higher median values in seronegative samples compared to seropositive samples for nucleocapsid IgG, RBD IgG, RBD IgA, and RBD IgM (p < 0.001 for each comparison, **Figure 4**). Seropositive samples were also more likely to have undetectable antigenemia. Similar trends were seen in the late-presenting group except for the comparison based on IgM which was not significant (**Supplementary Figure 7**).

**Figure 4.**
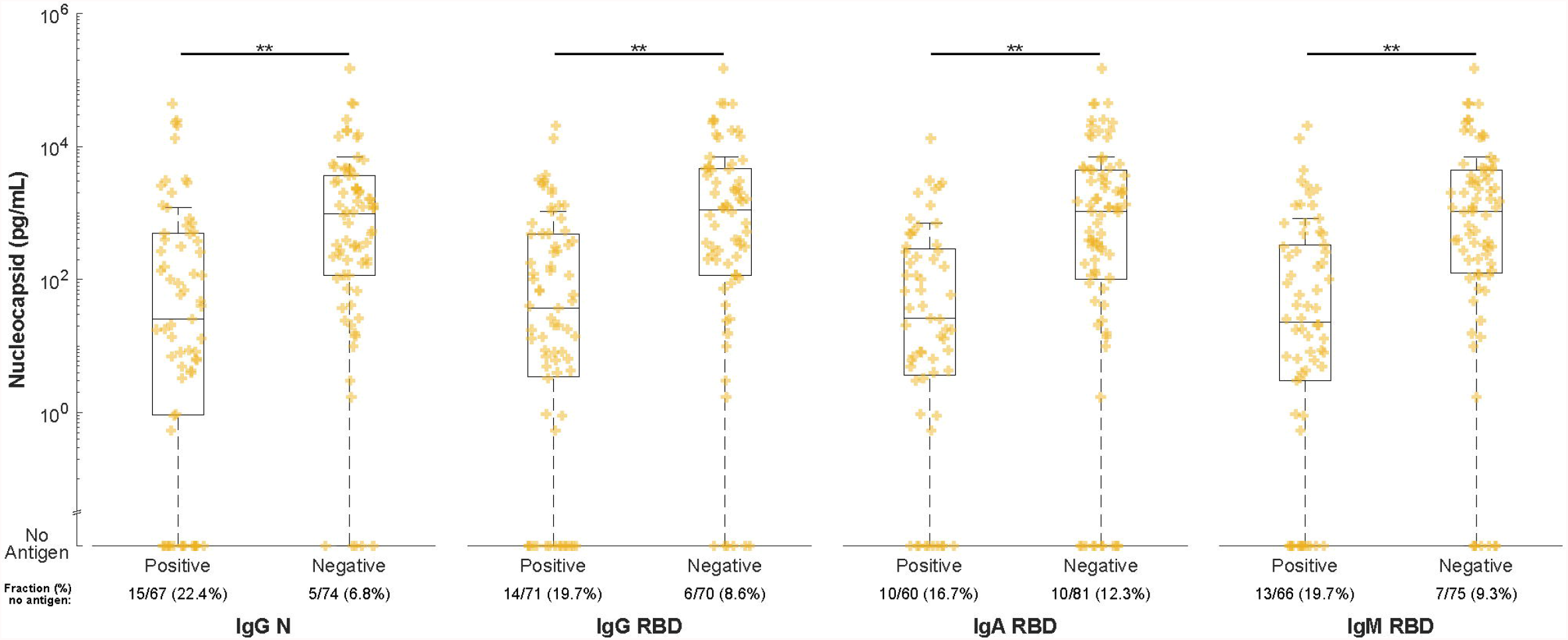
Comparison of serum or plasma nucleocapsid levels in individuals with and without SARS-CoV-2-specific antibodies. Samples were tested by in-house developed serological tests for nucleocapsid and receptor binding domain specific IgG as well as receptor binding domain specific IgA and IgM. Levels of nucleocapsid are plotted and compared in samples stratified by seropositivity for each antibody type.

### Association of Antigenemia with COVID-19 severity

In the acute COVID group, distribution of nucleocapsid antigen was significantly different and median value was higher in samples from patients who died or required intubation within 30 days of sampling compared to those who survived or did not require intubation (**Figure 5A-C**). This observation held true for comparison based on the composite of intubation or mortality. Levels of nucleocapsid antigenemia were not significantly associated with elevated D-dimer (cutoff 500 ng/mL) but were associated with elevated CRP (p = 0.002 in comparison based on 40 mg/L cutoff; **Figure 5D-E)**.

**Figure 5.**
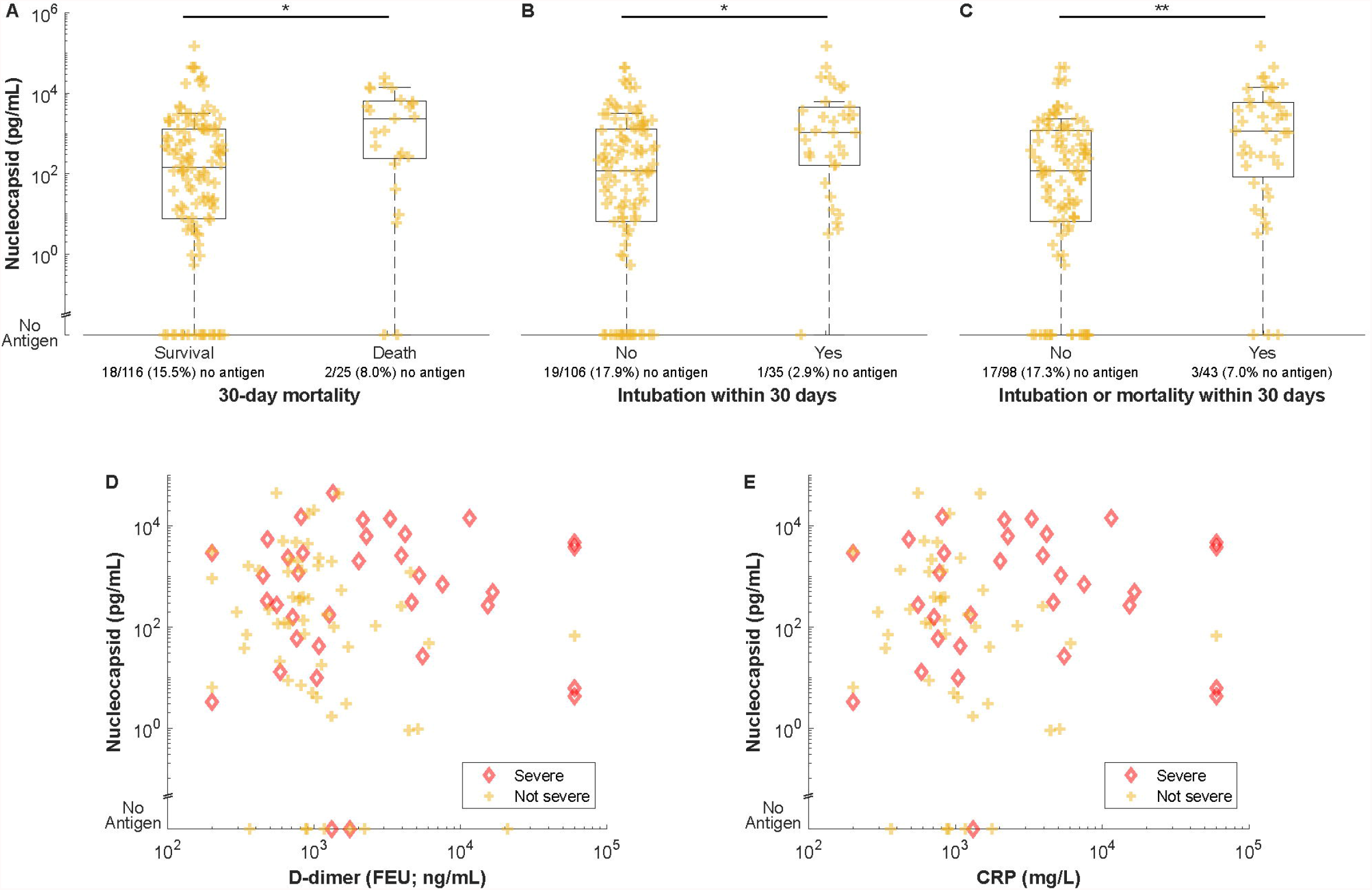
Comparison of serum or plasma nucleocapsid levels by (A-C) severity and (D-E) inflammatory biomarkers. Intubation in figures B and C includes intubation within 30 days before or after the blood sample was collected. Individuals with severe COVID as defined by the composite of 30-day intubation or mortality are highlighted in D and E.

## Discussion

This analysis of blood samples from routine clinical specimens collected during the ongoing COVID-19 pandemic demonstrates the following: First, antigenemia is a sensitive and specific marker for acute SARS-CoV-2 infection. Second, nucleocapsid is elevated in samples without evidence of anti-nucleocapsid (IgG) and anti-spike (IgG, IgM, and IgA) seroconversion. Third, antigenemia is associated with disease severity.

Evolving CDC isolation guidance during the COVID-19 pandemic reflects the difficulty of objectively defining resolution of SARS-CoV-2 infection. Underlying this is the persistence of RNA targets beyond a period during which the immunocompetent individual is reasonably believed to harbor replication-competent virus [1-3]. Meanwhile, persistence of replication-competent virus for months has been demonstrated by viral culture in immunocompromised hosts [24-28]. This creates a diagnostic dilemma when RT-PCR is persistently positive for weeks after diagnosis, when re-infection with SARS-CoV-2 is a consideration, or when encountering positive SARS-CoV-2 RT-PCR test results in an asymptomatic individual without history of prior objective diagnosis or prior COVID-like illness. Our data suggest assessment of nucleocapsid antigenemia may assist providers in making judgments in these scenarios.

Further, our data compels interest in whether antigenemia may provide direct evidence of active viral replication, which aid in evaluation of infectiousness or guide therapeutics at an individualized level. For example, antiviral agents are not likely to benefit a patient without active SARS-CoV-2 replication. Clinical trial data therefore may be confounded by failure to stratify patients according to such a marker, as late presenters after cessation of viral replication would likely fail to show benefit or may even suffer harm from investigatory antiviral agents. In fact, recent evidence emphasizes benefit of antivirals early in infection [29]. In showing its association with acute SARS-CoV-2 infection and characterizing outliers, our data suggest that nucleocapsid should be further investigated as a marker of viral activity, infectiousness, and a predictor of therapeutic response.

Strengths of our study include a diverse cohort that is among the largest in which nucleocapsid antigenemia has been quantified to date and rigorous assignment of COVID-19 status through medical record review. Prior studies restricting the definition of a positive case to no more than two weeks after symptom onset report sensitivities between 90.9% and 97.5% and specificities between 94.2% and 100% [15-20] (see **Supplementary Table 6**), and our data are consistent with these findings. Of further interest, our data revealed detectable antigen in 11 (2.1%) of the blood samples obtained in the primary serosurvey even though these patients were never screened with nasopharyngeal RT-PCR testing in our healthcare system. These represent likely infectious patients who may have had a missed SARS-CoV-2 diagnosis and suggest a potential role for antigenemia screening in a population for whom blood is already being sampled to complement existing infection control measures.

We also detected antigenemia in a small number of patients with subclinical SARS-CoV-2 infection. Individuals who test positive for SARS-CoV-2 without antecedent or subsequent COVID-like symptoms either represent shedding of replication-competent virus *during* subclinical disease or persistent RNA shedding *following* subclinical disease. While we corroborate previous findings that levels of antigenemia are associated with disease severity [19, 20], the presence of antigenemia in five asymptomatic individuals with SARS-CoV-2 demonstrates that antigenemia can also be present in subclinical infection. Despite the difficulty associated with identifying these cases, further investigation of the prevalence of antigenemia in acute asymptomatic infection is needed to clarify its role in screening broad populations.

This study is limited by use of a convenience sampling approach and retrospective data collection. Symptom onset as recorded in the medical record can be subjective and influenced by recall bias. Because of the ubiquity of community-based testing, SARS-CoV-2 diagnosis was documented prior to evaluation in our healthcare system for a subset of these patients and was only known to us when documented in the clinical narrative in addition to being subject to biases and imprecision. In addition, nucleocapsid-specific immunoglobulin may interfere with quantitation of antigenemia in individuals who have seroconverted although it is currently unknown whether total (Ig-bound and unbound) antigen or free (unbound only) antigen is a more meaningful clinical indicator. The primary analysis relies on the assumption that each subject is immunocompetent, that immunocompetent hosts have similar duration of acute COVID-19, and that there are no other confounding factors which may result in prolonged antigenemia. Recognizing these limitations, we performed a post-hoc investigation of outlier cases, which facilitated hypothesis generation regarding reasons for prolonged antigenemia such as reduced renal function, prolonged critical illness, and immune compromise (**Supplementary Tables 1-5 & 7**). Several studies have demonstrated high specificity of antigenemia by evaluation of pre-pandemic samples [15, 17, 20], suggesting many false positives in our study are likely to have active infection beyond the parameters for acute infection defined in our reference standard schema. This will be further clarified as more robust comparisons to viral culture, sgRNA, RT-PCR Ct value, and respiratory antigen testing can be achieved.

Together our data demonstrate that nucleocapsid antigenemia is a sensitive and specific biomarker of acute COVID-19 wherein COVID-19 status is defined by time since earliest positive testing and symptom onset. We conclude that nucleocapsid antigenemia is a promising candidate biomarker for active viral replication – the definition of which is the presence of replication-competent virus in a host – recognizing that the available evidence points to this being an individualized process that cannot be broadly defined based on a timeline. Further prospective studies with rigorous documentation of clinical course and correlation with viral culture and other potential biomarkers of viral replication are needed.

## Data Availability

All data produced in the present study are available upon reasonable request to the authors

## Acknowledgments

The authors thank Lisa Cole, Cecillitha J Williams, and Mark Meyers for assistance with specimen collection and Heather Jones for assistance with obtaining Ct values.

## Funding

This work was supported by funds from the Woodruff Health Sciences Center COVID-19 Center for Urgent Research Engagement (CURE), NIH 1 U54 CA260563-01: Immune regulation of COVID-19 infection in cancer and autoimmunity to JDR, NIH R01 HL138656 COVID-19 supplement to SRS, NIH R01 AI138646 supplement to NSS, NIH K24 AI114444 to NRG, and NIH Grants U54 EB027690 03 and UL1TR002378.

## Conflicts of Interest

C.A.R.’s institution has received funds to conduct clinical research unrelated to this manuscript from BioFire Inc, GSK, MedImmune, Micron, Janssen, Merck, Moderna, Novavax, PaxVax, Pfizer, Regeneron, Sanofi-Pasteur. She is co-inventor of patented RSV vaccine technology unrelated to this manuscript, which has been licensed to Meissa Vaccines, Inc.

All other authors report no conflicts of interest.

## Human Subjects Research

This study was approved by the Institutional Review Board of Emory University for the use of residual clinical specimens. Written informed consent by the patients was not required.

## Notes

### Competing Interest Statement

C.A.R. institution has received funds to conduct clinical research unrelated to this manuscript from BioFire Inc, GSK, MedImmune, Micron, Janssen, Merck, Moderna, Novavax, PaxVax, Pfizer, Regeneron, Sanofi-Pasteur. She is co-inventor of patented RSV vaccine technology unrelated to this manuscript, which has been licensed to Meissa Vaccines, Inc.
All other authors report no conflicts of interest.

