## Supplemental material for "Nucleocapsid antigenemia is a marker of acute SARS-CoV-2 infection"

**Supplementary Materials**

*Supplementary Tables*

*Supplementary Figures*

**Supplementary Methods**

*Nucleocapsid antibody ELISA*

For nucleocapsid antibody testing, 6x-histidine tagged recombinant nucleocapsid protein was produced in *E. coli* and purified by Ni-NTA chromatography before coating on high-binding ELISA plates at 1 µg/mL and 4°C overnight. ELISA plates were then blocked in PBS containing 1% bovine serum albumin (BSA) and 0.2% tween 20, before addition of patient samples pre-diluted at 1:500. After washing, anti-nucleocapsid IgG was detected using horseradish peroxidase (HRP)-conjugated anti-human IgG (Jackson Laboratory). Conjugate and sample dilutions were selected to optimize sensitivity and minimize background in control samples collected prior to the pandemic. ELISA optical density (OD) cutoffs for seroconversion were chosen by receiver operating characteristic (ROC) analysis with area under the curve for all assays greater than 0.95 in samples collected >14 days post symptom onset in PCR confirmed SARS-CoV-2 cases

*Assignment of COVID-19 status to blood samples*

Samples were categorized and labeled as follows. First, if the patient had *never tested positive* for SARS-CoV-2, a blood sample was labeled *negative* only if negative respiratory SARS-CoV-2 testing was recorded on the same day as the blood sample (**Figure 1A.1**). Blood samples with no record of positive SARS-CoV-2 testing and without same-day SARS-CoV-2 testing were excluded from analysis due to uncertainty of COVID-19 status at the time of blood sample collection.

Next, any patient who had *ever tested positive* for SARS-CoV-2 was categorized based on date of the earliest positive test (**Figure 1A.2**). While CDC guidelines for the public recommend isolation for ten days following diagnosis with resolution of symptoms [3], local healthcare guidelines recommend discontinuation of isolation after fourteen days in most cases which has also been supported in the literature [19]. Meanwhile, a three-day period has been proposed as the typical time between infection and symptom onset [19]. Based on this timeframe, a blood sample was categorized as *convalescent* and labeled *negative* if the patient had positive respiratory testing more than fourteen days prior to sample collection as the sample would have been collected in the period following cessation of AVR in most hosts. Otherwise, the sample was categorized as *in-window positive* if the earliest positive respiratory test was within the −14-to-+3 day window or *post-window positive* if the earliest positive respiratory test was more than three days after the blood sample was collected.

*Post-window positive* samples were then further adjudicated based on whether interim testing was available between the date of the blood sample and the date of the positive respiratory test. If an interim negative SARS-CoV-2 respiratory test was available, the sample was labeled *negative*. If no interim testing was available, then the status was considered *unknown* and the sample was excluded from analysis.

Finally, *in-window positive* samples were further adjudicated based on symptom onset (**Figure 1A.3**). The same −14-to-+3 day window was considered. A patient with symptom onset more than 14 days prior to the blood sample was further categorized as *late COVID* and labeled *negative* because even though earliest positive respiratory testing was in the window, time since symptom onset would place the blood sample in the period after which AVR is expected to have ceased. Meanwhile, if symptom onset was in the window, then the patient is expected to have AVR and was categorized as *acute COVID* and labeled *positive*. Finally, a patient testing positive in the window but with symptom onset more than 3 days after the positive test was excluded due to the uncertainty of this testing-symptom onset sequence.

*RT-PCR threshold cycle*

All threshold cycle (Ct) values were obtained directly from reports produced by the manufacturer’s software. Assays included the cobas® SARS-CoV-2 Test on the cobas® 6800 platform (Roche Diagnostics), the CDC 2019-Novel Coronavirus (2019-nCoV) Real-Time RT-PCR Diagnostic Panel or the TaqCheck™ SARS-CoV-2 Fast PCR Assay (Thermo Fisher) on the Applied Biosystems™ 7500 Real-Time PCR System (Thermo Fisher), and the Xpert® Xpress CoV-2/Flu/RSV on the GeneXpert platform (Cepheid).

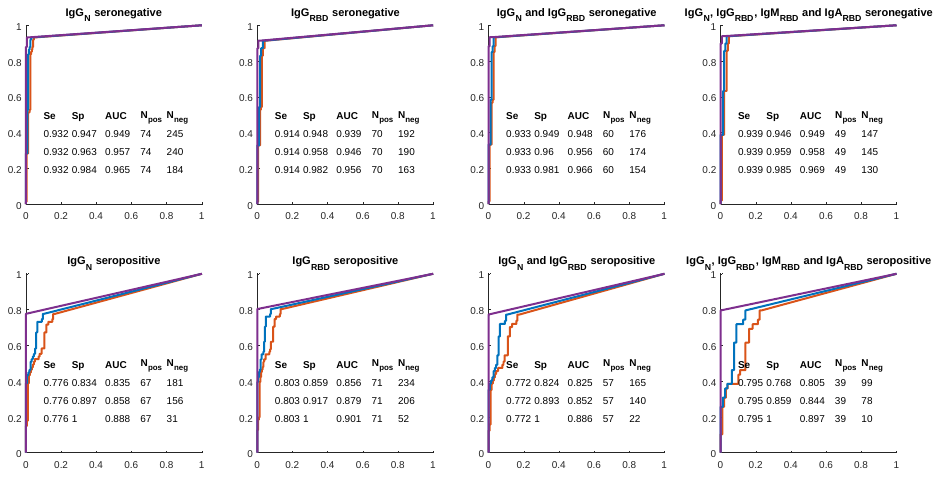

**Supplementary Figure 1.** ROC curves for nucleocapsid antigenemia within subgroups defined by serostatus.

Curves correspond to the same reference standards as Figure 2B in main manuscript (Red: acute vs all non-acute, Blue: acute vs convalescent/negative, Purple: acute vs negative only).

|  | **Age range** | **Days since COVID diagnosis** | **IgG N** | **IgG RBD** | **Mech vent** | **Death** | **Ct values** | **Clinical summary (at time of positive RT-PCR/negative antigenemia)** |
| --- | --- | --- | --- | --- | --- | --- | --- | --- |
| Sample  1050 | 71-80 | 39 | Yes | Yes | No | Yes | Roche 31.21 (ORF) 34.56 (E) | Presented 39 days after earliest positive test with non-respiratory illness |
| Sample  2047 | 61-70 | 29 | Yes | Yes | No | No | Roche 34.7 (ORF) E not detected | Pulmonary embolism > 1 week after resolution of primary COVID symptoms |
| Sample  255 | 71-80 | 21 | Yes | Yes | No | No | GeneXpert 35.2 | Presented with volume overload in the setting of missed dialysis |
| Sample  755 | 81-90 | 21 | No | Yes | No | No | Roche 32.47 (ORF) 33.84 (E) | Presented with abdominal pain, nausea and vomiting |
| Sample  2056 | 71-80 | 21 | Yes | Yes | No | No | CDC 34.69 (N1) 34.04 (N2) | Presented with hypoxic respiratory failure |
| Sample  141 | 41-50 | 16 | Yes | Yes | No | No | GeneXpert 30.9 | Presented with non-specific abdominal pain. No respiratory symptoms |
| Sample  995 | 61-70 | 15 | Yes | Yes | No | No | Roche  32.09 (ORF)  34.54 (E) | Presented with syncope |

**Supplementary Table 1.** Clinical details of patients with same-day positive SARS-CoV-2 nasopharyngeal NAAT and no antigenemia more than 14 days after diagnosis.

|  | **Days since first dx** | **Age range** | **Nucleocapsid (pg/mL)** | **Log_10_ Nucleocapsid (pg/mL)** | **IgG N** | **IgG RBD** | **Intubated** | **Death** | **Comment** | **Likely category** |
| --- | --- | --- | --- | --- | --- | --- | --- | --- | --- | --- |
| Samples 427* | 239 | 71-80 | 49.2 | 1.7 | Yes | Yes | No | No | Negative SARS-CoV-2 NP RT-PCR 12 days before, evidence of seroconversion in samples gathered 8 days apart. High risk for exposure to active cases. | Re-infection |
| and 150* | 231 | 71-80 | 2854.3 | 3.5 | No | No | No | No |  |  |
| Sample 121 | 130 | 41-50 | 1.9 | 0.3 | No | Yes | No | No | Recent high risk social gathering. | Re-infection |
| Sample 642 | 54 | 81-90 | 26.0 | 1.4 | No | No | No | No | Immunocompromised with COVID-like symptoms | Persistent infection in immunocompromised host |
| Sample 273 | 34 | 71-80 | 1.2 | 0.1 | Yes | Yes | Yes | Yes | Treated for hyper viscosity syndrome; same day blood transfusion | Severe COVID-19 |
| Sample 1352 | 33 | 71-80 | 19.7 | 1.3 | No | Yes | No | No | ESRD and recent chemotherapy; same day blood transfusion | Persistent infection in immunocompromised host |
| Sample 188 | 32 | 71-80 | 1.0 | 0.0 | Yes | Yes | No | No | Required high-flow oxygen after diagnosis. | Severe COVID-19 |
| Sample 2228 | 29 | 31-40 | 17.7 | 1.2 | Yes | Yes | Yes | No | ESRD; blood transfusion 4 days prior | Severe COVID-19 |
| Sample 1653 | 28 | 61-70 | 9.2 | 1.0 | Yes | Yes | Yes | Yes | Immunocompromised; same-day and prior day blood transfusion | Severe COVID-19 |
| Sample 683 | 26 | 61-70 | 0.9 | 0.0 | Yes | Yes | Yes | Yes | Immunocompromised | Severe COVID-19 |
| Sample 1067 | 20 | 41-50 | 2580.2 | 3.4 | No | No | No | No | ESRD, asymptomatic infection but admitted for diarrhea | Uncertain |
| Sample 2123 | 20 | 51-60 | 1.2 | 0.1 | Yes | Yes | Yes | No |  | Severe COVID-19 |
| Sample 1048 | 19 | 41-50 | 76.4 | 1.9 | Yes | Yes | Yes | No | Blood transfusion 3 days prior | Severe COVID-19 |
| Sample 2169 | 19 | 71-80 | 14.4 | 1.2 | Yes | No | No | Yes | Active malignancy; prior day blood transfusion | Uncertain |
| Sample 68 | 18 | 61-70 | 1.4 | 0.2 | Yes | Yes | No | No | Blood transfusion 4 days prior | Uncertain |
| Sample 1634 | 18 | 71-80 | 24.6 | 1.4 | Yes | Yes | No | No | Required high-flow oxygen | Uncertain |
| Sample 190 | 17 | 71-80 | 61.9 | 1.8 | Yes | Yes | No | Yes | ESRD; blood transfusion 3 days prior | Uncertain |
| Sample 841 | 17 | 71-80 | 652.7 | 2.8 | No | No | Yes | Yes | Blood transfusion 7 days prior | Severe COVID-19 |
| Sample 154 | 16 | 51-60 | 116.4 | 2.1 | Yes | Yes | Yes | No |  | Severe COVID-19 |
| Sample 1756 | 16 | 71-80 | 18.3 | 1.3 | Yes | Yes | Yes | Yes |  | Severe COVID-19 |
| Sample 651 | 16 | 71-80 | 0.4 | -0.4 | Yes | Yes | Yes | Yes |  | Severe COVID-19 |
| Sample 1992 | 16 | 71-80 | 5.0 | 0.7 | Yes | Yes | No | No | Diarrhea, not hypoxia | Uncertain |

**Supplementary Table 2.** Clinical details for samples in convalescent group with detectable antigenemia.

*Samples from the same patient. All other samples represent unique patients.

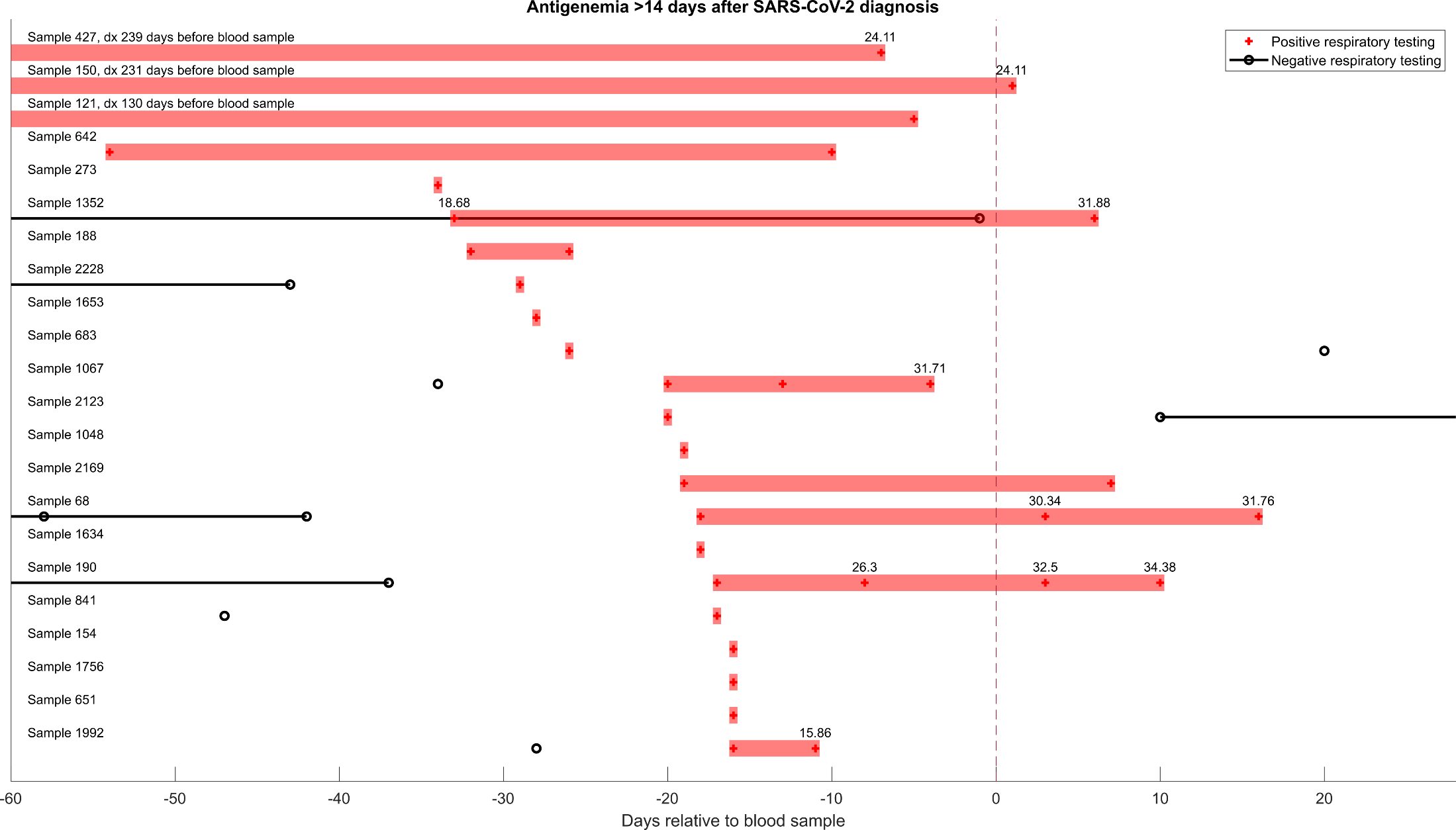

**Supplementary Figure 2.** Timeline of SARS-CoV-2 respiratory testing corresponding to samples with antigenemia in the convalescent group. Samples 427 and 150 are from the same individual. All other samples represent unique individuals. When available, Ct values are displayed above the markers for positive respiratory RT-PCR.

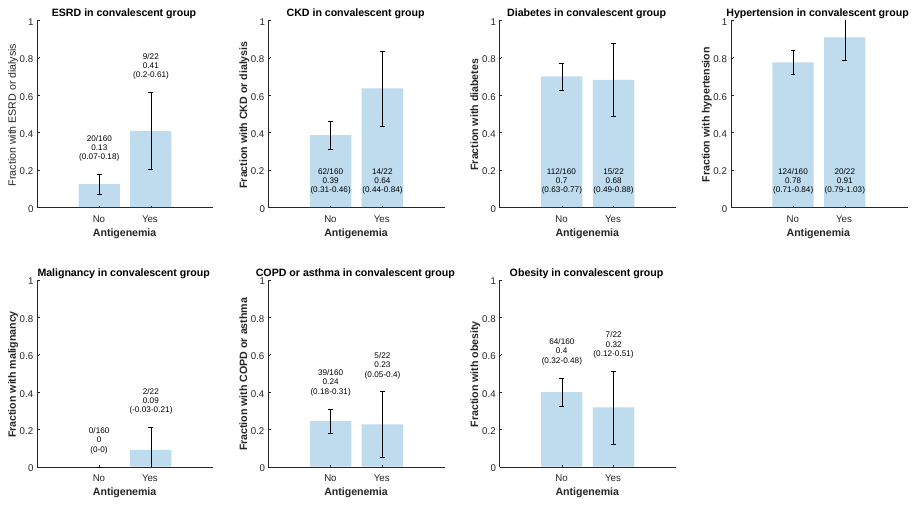

**Supplementary Figure 3.** Comparison of prevalence of comorbidities based on ICD10 codes for samples without and with antigenemia in the convalescent group. 95% confidence intervals are based on the standard error of the point estimate, i.e. $p\pm z\sqrt{\frac{p(1-p)}{n}}$ where $z$ = 1.96 and $n$ = sample size.

|  | **Age** | **N**  **protein** | **Log10**  **N protein** | **IgG**  **N** | **IgG**  **RBD** | **Comments** |
| --- | --- | --- | --- | --- | --- | --- |
| Sample 158 | 61-70 | 0.6 | -0.2 | No | No | Acute back pain.  No record of COVID-like symptoms. |
| Sample 1537 | 31-40 | 14.3 | 1.2 | No | No | Outpatient pre-op testing ahead of surgery.  No record of COVID-like symptoms. |
| Sample 1692 | 21-30 | 0.1 | -0.9 | No | No | Toxicology emergency.  No record of COVID-like symptoms. |

**Supplementary Table 3.** Same-day negative patients with antigenemia.

|  | **Age** | **Days since positive respiratory testing** | **Days since symptom onset** | **N protein (pg/mL)** | **Log10 N protein** | **IgG N** | **IgG RBD** | **Mechanical ventilation** | **Death** | **Quality of symptom documentation** |
| --- | --- | --- | --- | --- | --- | --- | --- | --- | --- | --- |
| Sample 2134 | 71-80 | 13 | 24 | 5.9 | 0.8 | Yes | Yes | Yes | No | Imprecise |
| Sample 540 | 81-90 | 12 | 19 | 27.0 | 1.4 | Yes | Yes | No | No | Imprecise |
| Sample 1354 | 61-70 | 6 | 19 | 19.1 | 1.3 | Yes | Yes | No | No | Imprecise |
| Sample 893 | 71-80 | 10 | 18 | 89332.6 | 5.0 | No | No | Yes | Yes | Imprecise |
| Sample 606 | 51-60 | 7 | 18 | 0.1 | -1.0 | Yes | Yes | No | No | Precise |
| Sample 280 | 61-70 | 5 | 18 | 3.9 | 0.6 | Yes | Yes | No | No | Imprecise |
| Sample 1702 | 21-30 | 14 | 17 | 11.9 | 1.1 | Yes | Yes | No | No | Precise |
| Sample 1574 | 81-90 | 14 | 17 | 1451.1 | 3.2 | Yes | Yes | No | Yes | Precise |
| Sample 1054 | 31-40 | 14 | 17 | 20.7 | 1.3 | Yes | Yes | Yes | No | Precise |
| Sample 221 | 71-80 | 14 | 16 | 21.0 | 1.3 | No | Yes | Yes | No | Imprecise |
| Sample 684 | 51-60 | 13 | 16 | 12.5 | 1.1 | Yes | Yes | Yes | Yes | Precise |
| Sample 167 | 71-80 | 9 | 16 | 45.4 | 1.7 | Yes | Yes | Yes | Yes | Imprecise |
| Sample 73 | 71-80 | 5 | 16 | 20.5 | 1.3 | Yes | Yes | No | No | Precise |
| Sample 690 | 21-30 | 14 | 15 | 3.9 | 0.6 | Yes | Yes | No | No | Precise |
| Sample 1725 | 71-80 | 14 | 15 | 773.6 | 2.9 | No | Yes | No | No | Precise |
| Sample 646 | 71-80 | 11 | 15 | 716.9 | 2.9 | No | No | Yes | Yes | Precise |
| Sample 179 | 71-80 | 8 | 15 | 942.0 | 3.0 | Yes | Yes | Yes | Yes | Imprecise |
| Sample 234 | 61-70 | 5 | 15 | 17.0 | 1.2 | Yes | Yes | No | No | Precise |

**Supplementary Table 4.** Patients with antigenemia and positive SARS-CoV-2 respiratory testing between 14 days prior to and 3 days after sample collection but with more than 14 days of symptoms at the time of sample collection. Quality of symptom documentation is categorized. Calendar dates (e.g. “anosmia started on Jan 1”) or integer-quantified history (e.g. “anosmia started 5 days ago”) are considered precise whereas other descriptions are considered imprecise (e.g. “anosmia a few days ago”).

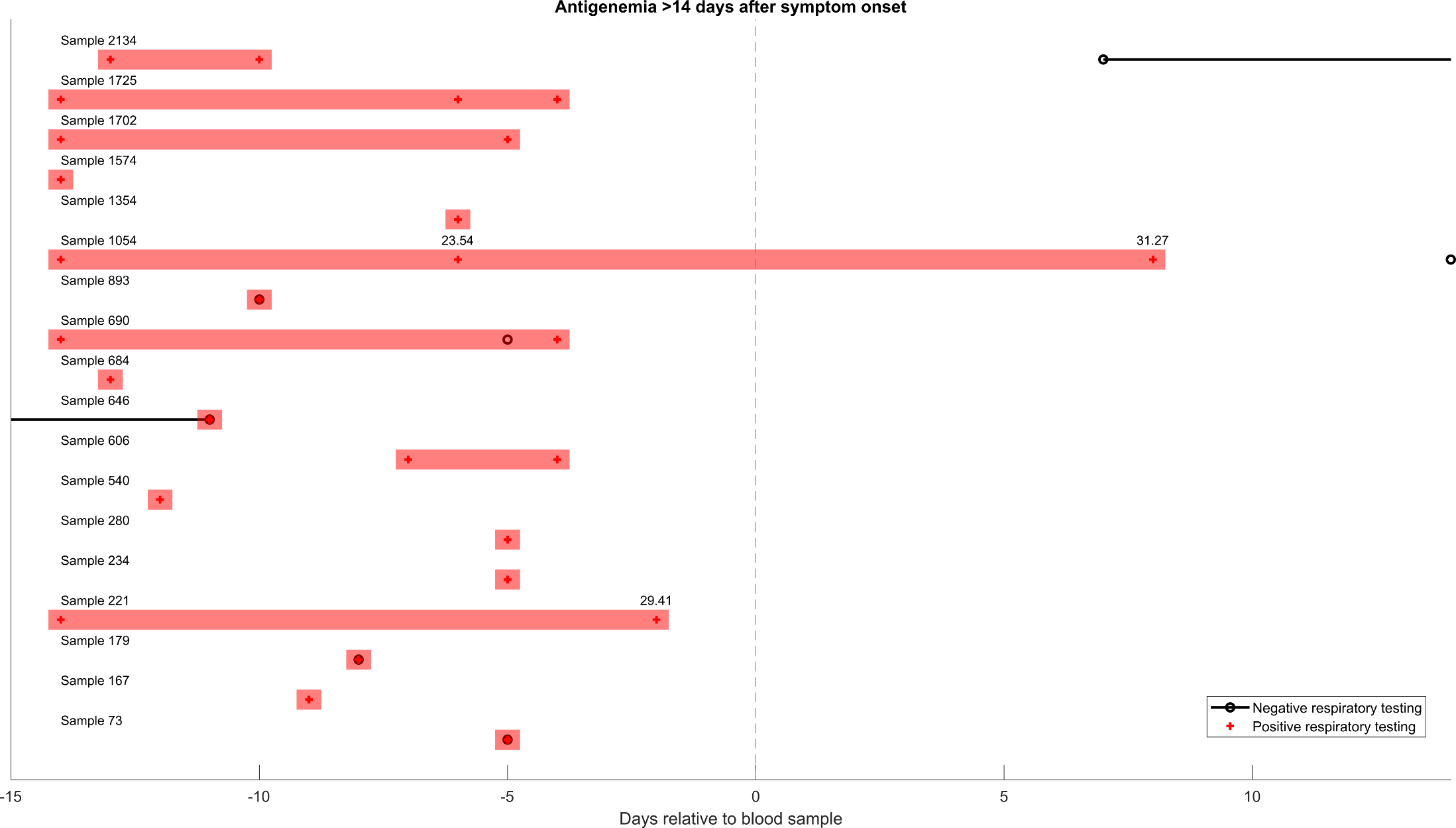

**Supplementary Figure 4.** Timeline of SARS-CoV-2 respiratory testing corresponding to samples with antigenemia in the late-presenting group (positive respiratory testing with 14 days prior to 3 days after the blood sample but with > 14 days of symptoms). When available, Ct value is indicated above positive tests.

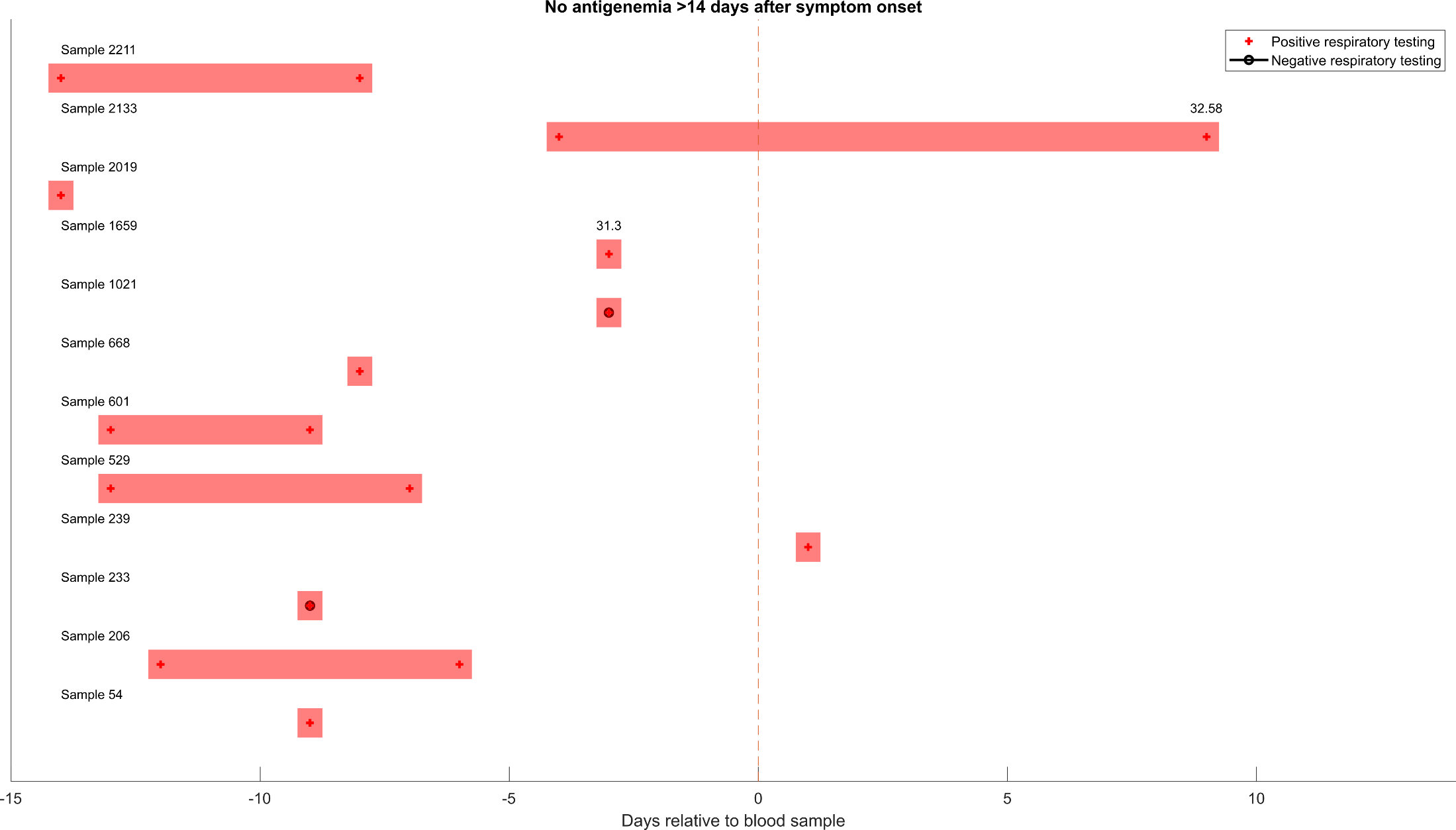

**Supplementary Figure 5.** Timeline of SARS-CoV-2 respiratory testing corresponding to samples without antigenemia in the late-presenting group (positive respiratory testing with 14 days prior to 3 days after the blood sample but with > 14 days of symptoms). When available, Ct value is indicated above positive tests.

|  | **Age** | **Days since**  **resp. test** | **Days since symptom onset** | **Ct value* (same day)** | **IgG N** | **IgG RBD** | **Mechanical ventilation** | **Death** | **Quality of symptom documentation** | **Comments** |
| --- | --- | --- | --- | --- | --- | --- | --- | --- | --- | --- |
| Sample 719 | 81-90 | 14 | 14 |  | Yes | Yes | No | Yes | Precise |  |
| Sample 1265 | 51-60 | 14 | 14 |  | No | Yes | No | No | Precise | Presented with fevers and chills. Reported to have had prior COVID diagnosis. |
| Sample 1045 | 51-60 | 7 | 7 |  | Yes | Yes | No | No |  |  |
| Sample 1878 | 61-70 | 14 | 14 |  | Yes | No | No | No | Precise | Reported prior positive testing with date not documented. Likely more than 14 days since first positive test. |
| Sample 119 | 31-40 | 7 | 14 |  | Yes | Yes | No | No | Imprecise |  |
| Sample 853 | 51-60 | 6 | 14 |  | Yes | Yes | No | No | Precise |  |
| Sample 122 | 71-80 | 7 | 13 |  | Yes | Yes | No | No | Imprecise |  |
| Sample 992 | 21-30 | 6 | 12 |  | Yes | Yes | No | No | Imprecise | No in-house testing available. |
| Sample 205 | 71-80 | 9 | 11 |  | Yes | Yes | No | No | Precise |  |
| Sample 602 | 41-50 | 12 | 10 |  | Yes | Yes | No | No | Imprecise |  |
| Sample 138 | 41-50 | 10 | 10 | 35.8 | Yes | Yes | No | No | Imprecise | Symptoms improving at time of blood sample. |
| Sample 1022 | 81-90 | 7 | 9 |  | No | No | No | Yes | Precise |  |
| Sample 421 | 71-80 | 3 | 8 |  | Yes | No | No | No | Precise |  |
| Sample 666 | 71-80 | 0 | 6 |  | Yes | Yes | No | No | Imprecise |  |
| Sample 970 | 71-80 | 4 | 5 |  | No | No | No | Yes | Imprecise | History provided by family member |
| Sample 738 | 21-30 | 3 | 5 |  | No | No | No | No | Precise |  |
| Sample 830 | 61-70 | 3 | 5 |  | No | No | No | No | Precise |  |
| Sample 1133 | 41-50 | 1 | 5 |  | Yes | Yes | No | No | Imprecise |  |
| Sample 365 | 51-60 | 1 | 1 |  | Yes | Yes | No | No | Imprecise | History provided by family member |
| Sample 760 | 71-80 | 0 | 0 |  | Yes | Yes | Yes | No | Imprecise | Presented with new cough. |

**Supplementary Table 5.** Samples categorized in the acute COVID group without antigenemia.

*if known

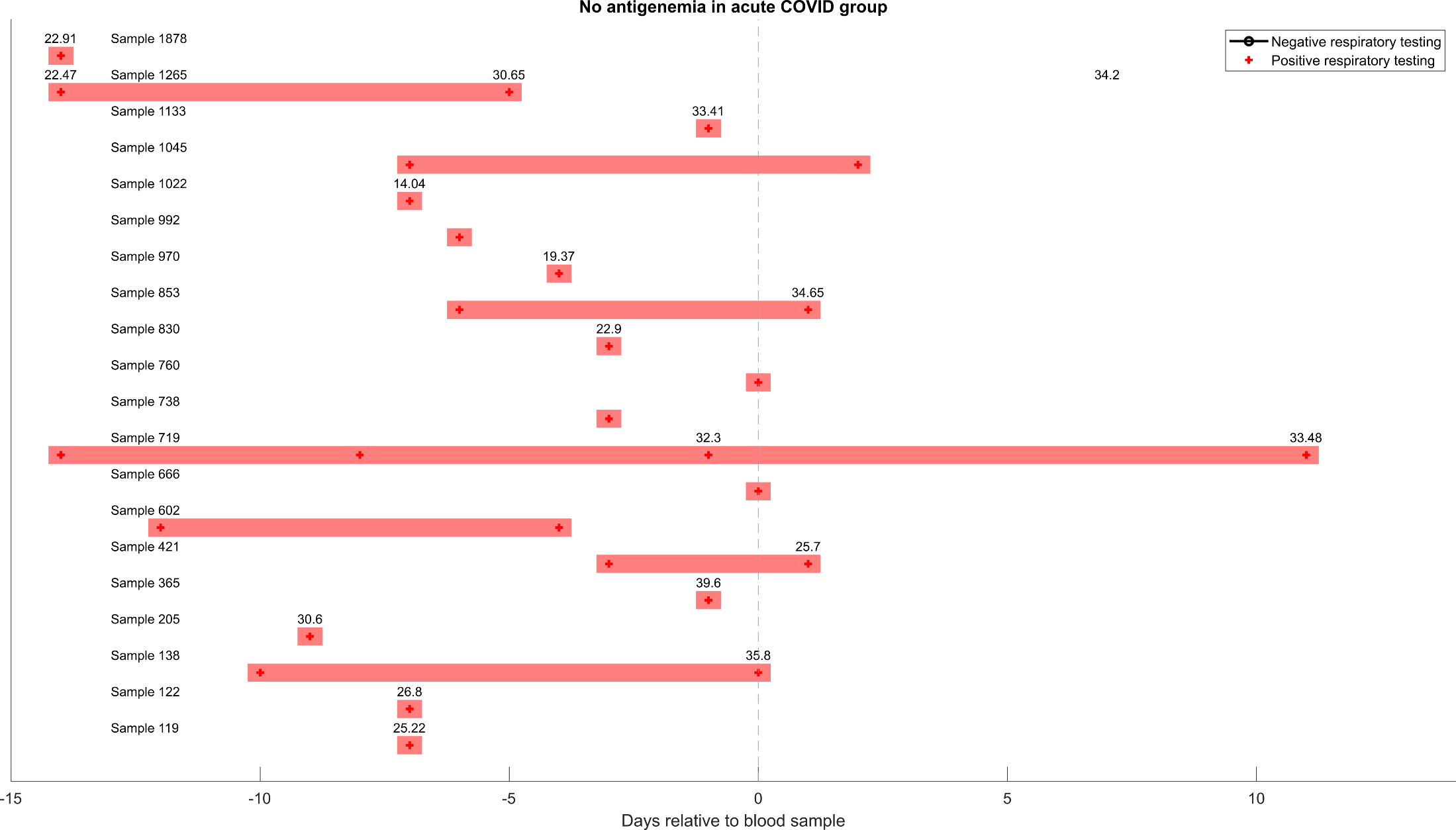

**Supplementary Figure 6.** Timeline of SARS-CoV-2 respiratory testing corresponding to samples without antigenemia in the acute COVID group (positive respiratory testing with 14 days prior to 3 days after the blood sample and < 14 days of symptoms). When available, Ct value is indicated above positive tests. Note that the patient corresponding to Sample 1265 had RT-PCR testing 7 days after blood sampling that was reported indeterminate (neither as positive nor negative) as only one of two targets amplified. Ct value for the target that did amplify appears here.

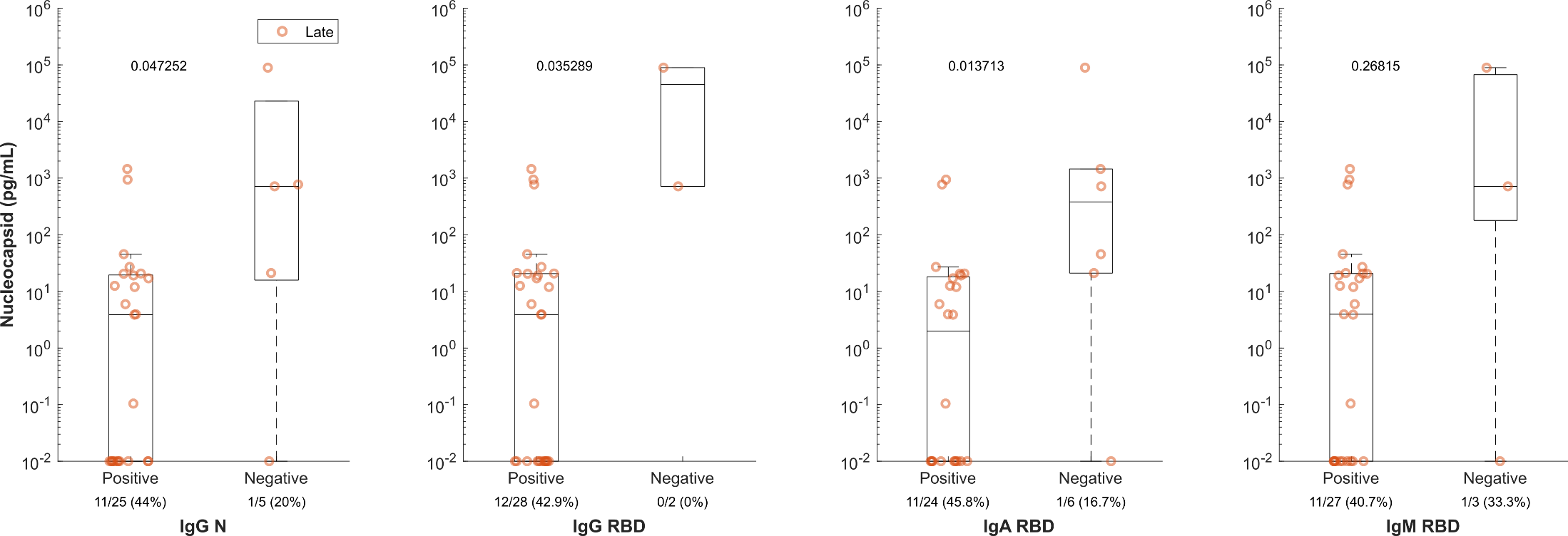

**Supplementary Figure 7.** Antigenemia levels in late-presenting group stratified by antibody serology.

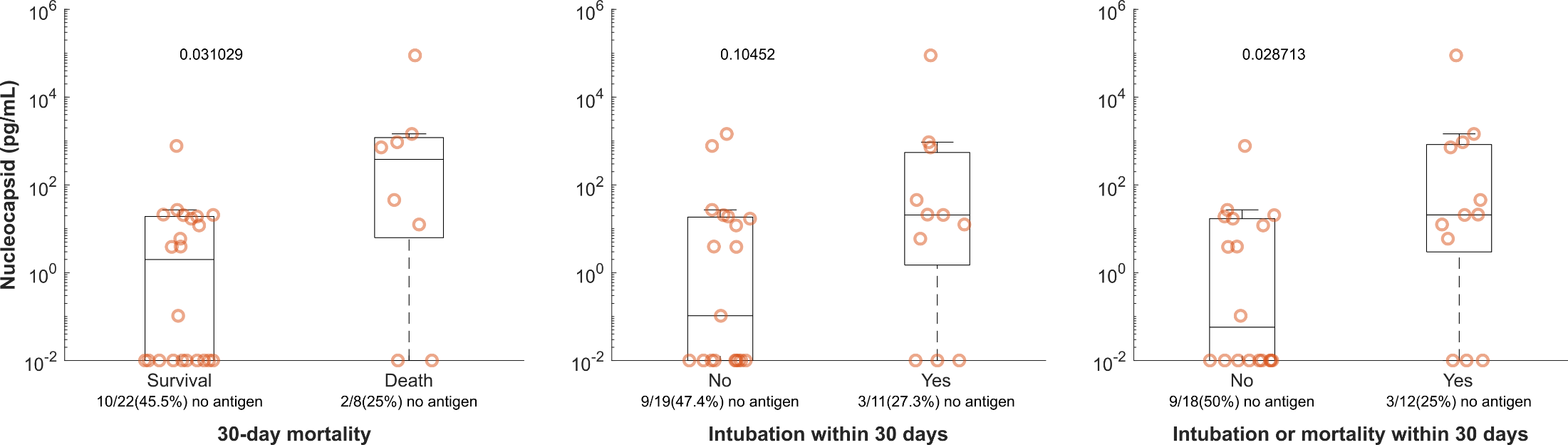

**Supplementary Figure 8.** Antigenemia levels in late COVID samples stratified by severity.

|  | Pub date | Assay | Design | Definition of COVID positive | Definition of COVID negative | Sens | Spec | Other data |
| --- | --- | --- | --- | --- | --- | --- | --- | --- |
| Li  Front Cell Infect Microbiol [11] | 9/4/2020 | ELISA | 40 COVID-19 patients from the First Affiliated Hospital of Anhui Medical University  and  30 COVID-19 patients from the Anhui Provincial Center for Disease Prevention and Control | Included patients had positive RT-PCR and pathological changes on chest CT, but test performance only reported in those who also did not have N-protein antibodies. | 633 with negative pharyngeal swab or sputum SARS-COV-2 nucleic acid result and negative N protein antibody (369 of these from patients with other respiratory infections) | 38/50 (76%) | 633/633  (100%) | Assay within-day & day-to-day precision |
| Lebedin [12]* | Pre-print posted 9/25/2020 | ELISA  (homebrew) | Patients with clinical signs of COVID-19 admitted to an redesigned facility | Unclear | Unclear |  | Unclear | ∙∙ |
| Su  Sci China Life Sci [13] | 11/26/2020 | Simoa  (Quanterix) | “89 plasma samples were obtained from 35 COVID-19 patients at different time points.” | Time from earliest positive RT-PCR positivity or symptom onset is not described. Supplemental information states: “Among 89 plasma samples, 39 were collected when qPCR assay of SARS-CoV-2 in pharyngeal swabs were positive.” | “healthy control” | 29/39  (74.4%) | 34/50  (68.0%) | ∙∙ |
| Ogata  Clin Chem  [14] | 11/30/2020 | Simoa  (Quanterix) | Adult patients presenting to Brigham and  Women’s Hospital or Massachusetts General Hospital | 10-days from initial NP RT-PCR test (does not account for symptom onset) | 17 RT-PCR negative  20 pre-pandemic healthy  14 pre-pandemic sick | 41/64  (64.1%) | NR | Clearance of plasma antigen  N positivity vs severity in Suppl. data  Multiple biomarkers (emphasis on S1 not N) |
| Hingrat [15] | 12/8/2020 | ELISA  (COVID-Quantigene) | Study participants included in the French COVID and CoV-CONTACT cohorts | Serum samples after SARS-CoV-2 diagnosis within 14 days of symptom onset. | Pre-pandemic samples and 13 pandemic samples with other viral infection | 132/142  (93.0%) | 62/63  (98.4%) | N vs RNAemia  N vs NP RT-PCR Ct |
| Ahava [16]* | Pre-print posted 1/13/2021 | ELISA  (Salofa) | Analysis of clinical samples sent to  Helsinki University Hospital Laboratory | Positive upper respiratory testing; blood sample within 14 days from symptom onset. | Samples from 2019 and 2020 | 47/50  (94.0%) | 145/148  (98.0%) | N vs Ct |
| Shan  Nat comm  [17] | 3/26/2021 | Simoa  (Quanterix) | SARS-CoV-2 RT-PCR+ samples from a commercial source (N = 20)  and  Longitudinal plasma from the University of Bonn (N = 20; symptom onset not recorded) | RT-PCR+; test characteristics are calculated from “first draw” sample data presented in supplement and based on cut-off of 1.25 pg/mL. Sample N according to days from positive PCR as follows:  1-7 days: N = 27  8-14 days: N = 12  > 14 days: N = 1 | Specificity is based on measurements of pre-pandemic samples. | 39/40  (97.5%) | 100/100  (100%) | Dried blood spots  Report antigenemia > 14 days in longitudinal cohort (53 samples) but *all from six of ten total patients* |
| Zhang  Clin Chem  [18] | 8/6/2021 | ELISA  (Biohit Healthcare) | Remnant serum from 208 randomly selected COVID cases at Zuckerberg San Francisco General Hospital. | RT-PCR positive within 24 hours, within 1 week of symptom onset. | RT-PCR negative | 130/143  (90.9%) | 59/60  (98.3%) | Absence of cross-reactivity for 5 individuals with other coronavirus infections |
| Thudium [21] | 9/20/2021 | ELISA  (Solsten Diagnostics) | Inpatients and outpatients | Confirmatory PCR-positive test within 13 days | Simultaneous upper respiratory RT-PCR negative | 282/324  (87.0%) | 1/462  (99.8%) | Diagnostic cutoff value of 10 pg/ml NP |
| Wang  Clin Chem  [19] | 10/4/2021 | S-PLEX Direct Detection Assay  (Meso Scale) | Remnant venipunctures from patients receiving blood draws between March to November 2020 at Stanford Healthcare. | Blood samples +/- 1 day from first positive NAAT, sensitivity summarized here are for those tested within 2 weeks of symptom onset. | SARS-CoV-2 NAAT negative | 64/70  (91.4%) | 49/52  (94.2%) | Comparison to Ct values  Severity analysis |
| Favresse [20]* | Pre-print posted 11/21/2021 | Simoa  (Quanterix) | All patients with molecular diagnosis of SARS-CoV-2 infection between April 2020 and July 2021. | Within 10 days of symptom onset. | 71 pre-pandemic serum samples collected before February 2020. | 93/96  (96.9%) | 70/71  (98.6%) | Severity analysis  Also presents iFlash assay data |

**Supplementary Table 6.** Summary of current literature

*preprint; NR = not reported.

|  | **Convalescent** | **Late COVID** | **Acute COVID** | **Pre-COVID** | **Same-day negative** |
| --- | --- | --- | --- | --- | --- |
| **COVID status label** | Negative | Negative | Positive | Negative | Negative |
| **Earliest positive SARS-CoV-2 respiratory testing** | > 14 days before blood sample | 14 days before to 3 days after blood sample | 14 days before to 3 days after blood sample | > 3 days after blood sample | Never |
| **Negative SARS-CoV-2 respiratory testing** | N/A | N/A | N/A | After blood sample and before earliest positive test | Same day as blood sample |
| **Symptom onset** | N/A | > 14 days before blood sample | 14 days before to 3 days after blood sample | N/A | N/A |
| **Proposed reasons for unexpected presence of antigenemia** | Re-infection  Critical illness (prolonged infection)  Immune compromise (chronic infection) | Critical illness (prolonged infection)  Immune compromise (chronic infection)  Decreased antigen clearance (ESRD, CKD or AKI) | N/A | Undiagnosed COVID at time of blood sample with late respiratory testing | False-negative SARS-CoV-2 respiratory testing |
| **Proposed reasons for unexpected absence of antigenemia** | N/A | N/A | Late in acute period (~7-14 days after symptom onset).  Imprecise or inaccurate symptom history obtained from retrospective chart review.  Earliest positive SARS-CoV-2 test more than 14 days prior but not documented.  Persistent SARS-CoV-2 RNA following subclinical acute infection and now presenting with symptoms related to another process | N/A | N/A |

**Supplementary Table 7.** Outline of blood sample classification scheme and discussion of proposed reasons for variation from the antigenemia as a unique and universal marker of acute infection
